# Is mandatory vaccination in population over 60 adequate to control the COVID-19 pandemic in E.U.?

**DOI:** 10.1101/2022.01.25.22269867

**Authors:** N.P. Rachaniotis, T.K. Dasaklis, F. Fotopoulos, M. Chouzouris, V. Sypsa, A. Lyberaki, P. Tinios

## Abstract

Vaccine hesitancy, which potentially leads to refusal or delayed acceptance of COVID-19 vaccines, is considered a key driver for the increasing death toll from the pandemic in the E.U.. European Commission and several member states’ governments are either planning or have already directly or indirectly announced mandatory vaccination for individuals aged over 60, the group repeatedly proved to be the most vulnerable. In this paper, an assessment of this strategy benefits is attempted. This is done by examining the reduction of Standard Expected Years of Life Lost (SEYLL) per person of the EU population over 60 as a function of their vaccination percentage. Publicly available data and some first results of the second iteration of the SHARE COVID-19 survey conducted during the summer of 2021 on acceptance of COVID-19 vaccines are used as input.

## Introduction

Two years after its emergence in Wuhan, China, the global spread of SARS-CoV-2, the pathogen that caused the pandemic COVID-19, continues to strain healthcare systems. By January 3^rd^, 2022, 290,157,607 confirmed cases of COVID-19, including 5,443,753 deaths, were reported globally (COVID-19 Map - Johns Hopkins Coronavirus Resource Center (jhu.edu)). Up to December 2020, due to the absence of effective therapies and vaccines, countries were limited to Non-Pharmaceutical Interventions (NPIs) to control the spread of the disease and minimize death rates. They included, inter alia, social distancing and lockdowns, travel restrictions, face masks, teleworking, etc. As vaccines became available, countries tried to develop exit strategies from the pandemic. These strategies aimed to balance the optimum utilization of vaccine resources which became gradually available) and the retention of some NPIs.

Over the same period, the European Union (E.U.) reported more than 57,000,000 confirmed cases and 906,000 deaths (COVID-19 Data Explorer - Our World in Data). The increasing death toll from the pandemic in E.U. was attributed to two major reasons: vaccine hesitancy, which lies behind refusal or delayed acceptance of vaccines, and the advent of the Delta variant in March 2021. The latter substantially increased the waning of vaccines’ effectiveness, especially for ages over 60 – those who account for more than 90% of the total deaths from COVID-19 in Europe (Hoffmann and Wolf, 2021).

The consequence is an increasing discussion across the E.U. of ways to convince the unvaccinated. With the notable exception of health care personnel, who were obliged to get vaccinated in several E.U. countries until November 2021, the measures relied on negative incentives, in the form of COVID-19 “green passes” to limitations targeting only the unvaccinated population. However, an increasing number of countries have implemented or consider implementing mandatory vaccination (Table 1), while the European Commission has called for a horizontal E.U. discussion on the topic. Vaccination is defined as a legal obligation for sub-sections of the population, while direct penalties are envisaged in the form of fines or suspension of employment. However, even where the threat of mandatory vaccination exists, a significant fraction of the population remain unconvinced and appear to deny vaccination at all costs.

How can we judge the potential efficacy of vaccine mandates? This question necessitates computing a counterfactual. This would rest on assuming how many more people will be vaccinated as a *direct* result of the mandate, who would not have been vaccinated otherwise. This assumption needs to be supplemented by a metric of the gains from vaccination. This paper chooses for this purpose the Standard Expected Years of Life Lost (SEYLL) per person of E.U. population over 60. SEYLL is a standard metric used in burden-of-disease estimations. It calculates life years lost compared with a standardised reference life table -which in the case examined is common across the EU. In other words, the gains of the mandates are identified as the *averted* SEYLL should the mandated obligation succeed – i.e. if the people who would not have been vaccinated on their own are pushed to be protected.

**Table 1.**
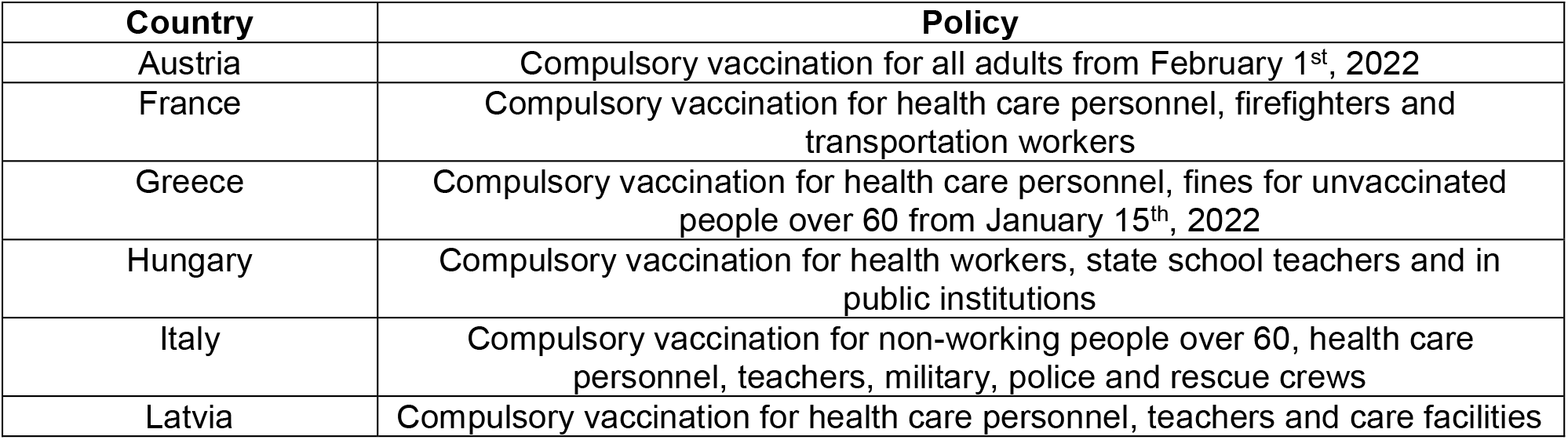
E.U. countries compulsory vaccination policies by January 6, 2022.

Publicly available data are used to compute the benchmark SEYLL, while findings from the second iteration of the SHARE COVID-19 survey conducted during the summer of 2021 are used to proxy the numbers of people in each country who resist voluntary vaccination.

The results showcase policy recommendations regarding the pandemic containment in E.U.. They could support decision-makers in suggesting a policy pathway to be followed until the majority of the population is vaccinated and argue in favor or against vaccine mandates at a country or E.U. level.

The remainder of this paper is organized as follows. Section 2 provides a review of the relevant literature. In Section 3, the proposed methodology of the study is described, whereas in Section 4 the results are presented. Finally, Section 5 discusses the study’s results and their policy implications.

## Background literature

Large differences in country-specific COVID-19 mortality rates have prompted debate and speculations about the reasons that lie behind it. Many factors have been examined recently, including genetic, viral, medical, socioeconomic and environmental ones. Nevertheless, reliable data proved from the very beginning that the crude Case Fatality Rate (CFR) of COVID-19 is predominantly determined by the proportion of population aged older than 60 years of all individuals diagnosed with SARS-CoV-2 infection (Hoffmann and Wolf, 2021). In consequence, the question that naturally arises is whether a targeted vaccination of the ages over 60 would be sufficient in order to mitigate the impact of the pandemic.

Trying to answer this question, a literature stream, even before the advent of vaccines, focused on the possible benefits of combining vaccination with NPIs with various levels of vaccines efficiency (Getz et al., 2020; Choi and Shim, 2020; Iboi et al., 2020; Maltsev and Stern, 2020). In order to eliminate disease transmission, a highly effective vaccine is required and complementing vaccination with NPIs will always yield optimal containment results (Kuzdeuov et al., 2020).

Vaccination against COVID-19 initiated globally in December 2020. After a year of continuous testing, it is proven that vaccine effectiveness against symptomatic disease and hospitalization falls significantly after a time interval of 20-25 weeks (5 to 6 months) from their vaccination completion against the dominating till December 2021 Delta variant. Waning is faster in older adults and those in a clinical risk group (Andrews et al., 2021; Han et al., 2021).

Vaccines aim to reduce premature deaths caused directly or indirectly by COVID-19. To capture vaccines efficiency in this paper, the metric used is SEYLL per living person aged over 60. SEYLL, a specific form of Years of Life Lost (YLL), is a standard metric used in burden-of-disease estimations. It calculates life years lost compared with a standardised reference life table. In other words a person’s life expectancy at each age is estimated based on the lowest observed age-specific mortality rates *across all countries in the EU*. The major features of this choice are:

- It regards all deaths as important but those affecting younger individuals (but still over 60) as particularly important, given that for their case more life years are lost. This has the indirect impact of placing greater weight on lower income E.U. countries in which younger deaths are more common. Thus, understanding the mortality impact of COVID-19 requires not only counting the dead but also analyzing how premature those deaths are
- It values a death at a given age identically across all countries in the EU, regardless of differences in national life expectancy or income per capita. Otherwise a death occurring in a high longevity, richer country would count for more than one occurring in a poorer, less long lived country.

Given its egalitarian emphasis, this metric is selected in the proposed Fair Priority Model for allocation of vaccines, thus rendering it a suitable assessment tool (Emanuel et al., 2020; Herzog et al., 2021; Wouters et al., 2021).

YLL have been used to assess COVID-19 effects in some papers, either studying a single country (e.g. Quast et al., 2020; Rommel et al., 2021) or several countries together(e.g. Mitra et al., 2020; Oh et al., 2020; i Arolas et al., 2021; Vieira et al., 2021) – though before vaccination was available. Only Ferenci’s (2021) research work took place in the first months of the vaccination campaign in Hungary.

This paper attempts to express SEYLL from COVID-19 per living person aged over 60 in E.U. as a function of population vaccination coverage. Developing such an aggregated model could be useful for policymakers to assess strategies that favor mandatory vaccination affecting this age group, both at a country or even at E.U. level, in terms of how much these strategies can reduce the total death toll should a particular vaccination level can be achieved.

## Methodology

Available country-level age- and gender-specific data on COVID-19 deaths for the ages over 60 in the 27 E.U. countries from 25^th^ May 2021 to 25th November 2021 from the COVerAGE database (Riffe et al., 2021), freely available from the Open Science Framework (Riffe et al., 2021), were used. The COVerAGE data was processed and stratified to three 10-year intervals, i.e. 60-69, 70-79 and 80+, separately for females and males, across countries. Countries’ missing data were filled in from their national health authority websites and national press reports. Age-stratified data to the same 10-year intervals on weekly numbers of vaccinations in E.U. countries up to the 25^th^ of November 2021 were collected from the European Centre for Disease Prevention and Control (ECDC). Germany and Netherlands that do not provide these data, so were filled in from national press reports and relevant research work filled in.

This specific six-months period (25/5/2021-25/11/2021) was selected for three reasons:

1. The majority of E.U countries were able to provide their citizens over 60 the opportunity to complete their vaccination until the end of May 2021
2. Vaccines effectiveness wanes significantly after 6 months from its completion, especially among the elderly.
3. On November 26^th^ 2021 the WHO classified Omicron as a variant of global concern, which eventually prevailed over Delta variant at the end of December in most E.U. countries. The Omicron variant is still under examination regarding its CFR and vaccines effectiveness against it. Therefore it is not possible yet to gauge the mandatory vaccination benefits for the 60+ age group.

The E.U. population over 60 intending to be vaccinated was captured from the Survey of Health, Ageing and Retirement in Europe (SHARE) (www.share-project.org). The analysis used data from the second iteration of the SHARE COVID-19 Survey. This re-interviewed 46,989 respondents aged 50 years and older (91% of whom were 60 years and older) of the first SHARE COVID-19 Survey, and was fielded from June to August 2021 in all 28 countries participating in SHARE (26 EU countries plus Switzerland and Israel). SHARE’s second COVID-19 survey remains one of the few, if not the only large-scale study, that covers all E.U. countries and has collected data on individuals’ situation during the pandemic, using sampling methodologies that provide internationally comparable data.

All respondents were asked whether they had been vaccinated against COVID-19 at least once. *“Information on their intention to do so was requested—inquiring whether they had already scheduled an appointment for vaccination, wanted to get vaccinated, were still undecided or did not want to get vaccinated at all”* (Bergmann et al., 2021). It is this last percentage – i.e. those adamant they would not be vaccinated – that we use to proxy the *upper* vaccination limit in case of fully enforcing mandatory vaccination for the ages over 60 for every E.U. country in SHARE. Given that the mandatory vaccination strategy is considered predominately in the EU, the two non-EU countries, Switzerland and Israel, were excluded from the analysis.

The total expected years of life lost due to COVID-19 for the over-60 population, denoted SEYLL_t_, is

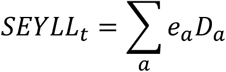

where:

D_a_: denotes deaths in the six-month period due to COVID-19 in a 10-years interval a, a=“60-69”, “70-79”, “80+”.

e_a_: is a measure of the expected years of life that remain to be lived for a death from any cause at age a. These coefficients are specified by MLTW tables, separately for females and for males, thus forming the ‘‘standard’’ in the SEYLL metric.

In order to relate SEYLL_t_ to the population structure of E.U. countries and to capture their significanty different mortality rates, the years lost per death SEYLL_d_ and the years lost per living person SEYLL_p_ are considered (Marshall, 2009):

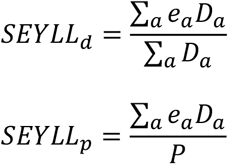

where P: is the population size for people over 60. E.U. countries’ population structure projection for 2021 by age and gender (www.ec.europa.eu/eurostat) is used.

## Results

Table 2 illustrates the actual vaccination percentages by 25/11, all of which predated mandatory vaccinations, and are hence fully voluntary. The table shows SEYLL_d_ in years and SEYLL_p_ in days from 25/5 to 25/11 for the 60+ age group in the 27 E.U countries. The calculated SEYLL_d_ values (mean=11.3 years, s.d.=0.74 years) – shown in Figure 1 - are consistent with the results of several relevant studies, where SEYLL_d_ were estimated between 10 and 13 years in developed countries (Hanlon et al., 2020). The metric is useful for comparison purposes to other causes of death *within* E.U. countries, e.g. in most of them COVID19 is the third cause of death after cancer and cardiovascular diseases (Vieira et al, 2021). However, it is not capturing the significantly different mortality *rates* (deaths per million people) *between* countries. This is obtained with the calculation of SEYLL_p_.

**Table 2.** Vaccination percentages by 25/11, SEYLL_d_ in years, SEYLL_p_ in days from 25/5 to 25/11 for the 60+ age group, upper vaccination limits for 60+ age group and estimated SEYLL_p_ for the 27 E.U. countries.

| Country | Voluntary Vaccination percentage by 25/11 in the 60+ age group | SEYLL<sub>d</sub> (in years) | SEYLL<sub>p</sub> (in days) for the 60+ age group from 25/5 to 25/11 | Upper vaccination limit from SHARE COVID-19 survey (%) for ages 60+ | SEYLL<sub>p</sub> estimation (days) |
| --- | --- | --- | --- | --- | --- |
| Austria | 87.9 | 11.40 | 1.40 | 90.51 | 1.58 |
| Belgium | 93.6 | 11.37 | 1.43 | 97.65 | 0.64 |
| Bulgaria | 32.9 | 10.74 | 9.19 | 63.30 | 5.14 |
| Croatia | 70.8 | 11.54 | 4.98 | 90.43 | 1.59 |
| Cyprus | 92.6 | 12.81 | 3.29 | 93.60 | 1.17 |
| Czechia | 82.3 | 10.65 | 1.79 | 93.11 | 1.24 |
| Denmark | 99.1 | 10.71 | 0.44 | 99.10* | 0.45 |
| Estonia | 72.8 | 12.22 | 3.62 | 90.07 | 1.63 |
| Finland | 93.3 | 10.70 | 0.44 | 97.84 | 0.62 |
| France | 85.7 | 12.70 | 1.42 | 94.62 | 1.04 |
| Germany | 90 | 10.79 | 0.76 | 95.29 | 0.95 |
| Greece | 79.6 | 11.21 | 3.84 | 92.38 | 1.33 |
| Hungary | 80.1 | 10.73 | 2.88 | 92.67 | 1.29 |
| Ireland | 100 | 11.86 | 1.80 | 100.00 | 0.33 |
| Italy | 89.4 | 10.72 | 1.15 | 96.93 | 0.74 |
| Latvia | 65.5 | 12.62 | 5.30 | 77.69 | 3.25 |
| Lithuania | 75.2 | 10.05 | 6.34 | 84.00 | 2.43 |
| Luxembourg | 86.8 | 10.32 | 0.93 | 94.96 | 0.99 |
| Malta | 98.8 | 11.16 | 0.81 | 98.80 | 0.49 |
| Netherlands | 94 | 12.08 | 0.80 | 97.43 | 0.67 |
| Poland | 74.7 | 10.70 | 1.88 | 89.67 | 1.68 |
| Portugal | 99.5 | 11.80 | 0.84 | 99.50* | 0.40 |
| Romania | 42.4 | 10.73 | 8.85 | 63.81 | 5.07 |
| Slovakia | 67.5 | 11.17 | 2.23 | 86.02 | 2.16 |
| Slovenia | 80.2 | 11.17 | 2.95 | 88.97 | 1.78 |
| Spain | 97.5 | 11.32 | 1.29 | 97.82 | 0.62 |
| Sweden | 93.1 | 11.47 | 0.55 | 99.17 | 0.44 |

\* Actual vaccination percentages by 25/11/2021

**Figure 1.**
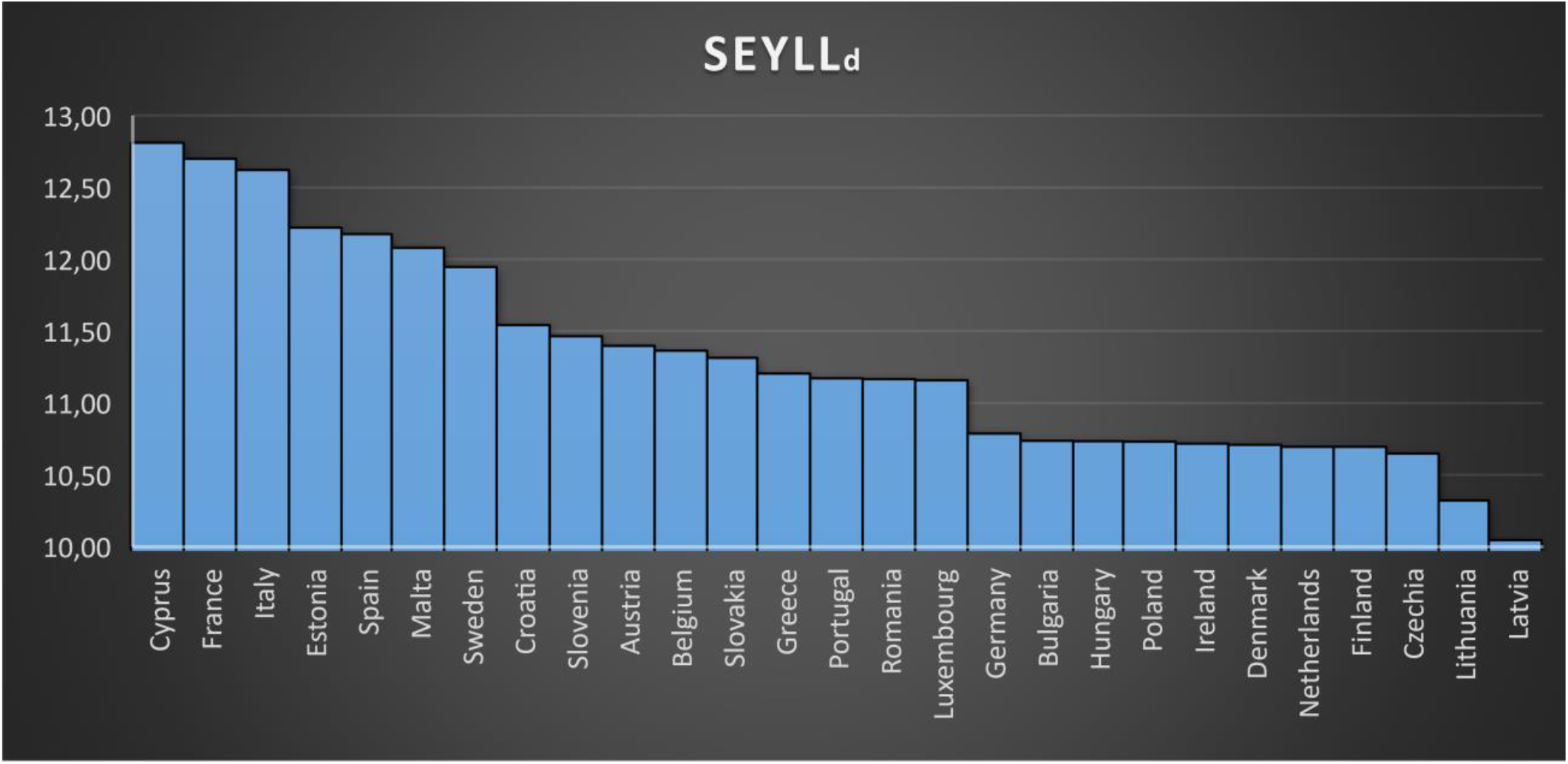
SEYLL_d_ for the 27 E.U. countries.

The days of life lost *per living person aged over 60* for the examined 6-months period (mean=2.64 days, s.d.=2.42 days, min=0.44 days, max=9.19) form a representative metric of the significant pandemic’s negative effects on every E.U. country. They measure how many fewer days a representative individual over 60 expectσ to live, as a direct result of COVID-19 morbidity occurring in these six months.

In order to relate SEYLL_p_ for the population over 60 to their vaccination percentage coverage, an approximated value function was calculated from the data of Table 1 using the least squares method. The best approximation function calculated was (Figure 2):

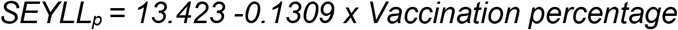

with R^2^=79.62%. The 95% CI for the intercept is [11.13, 15.71] (s.e. = 1.11) and for the slope is [- 0.16, -0.10] (s.e.= 0.01).

**Figure 2.**
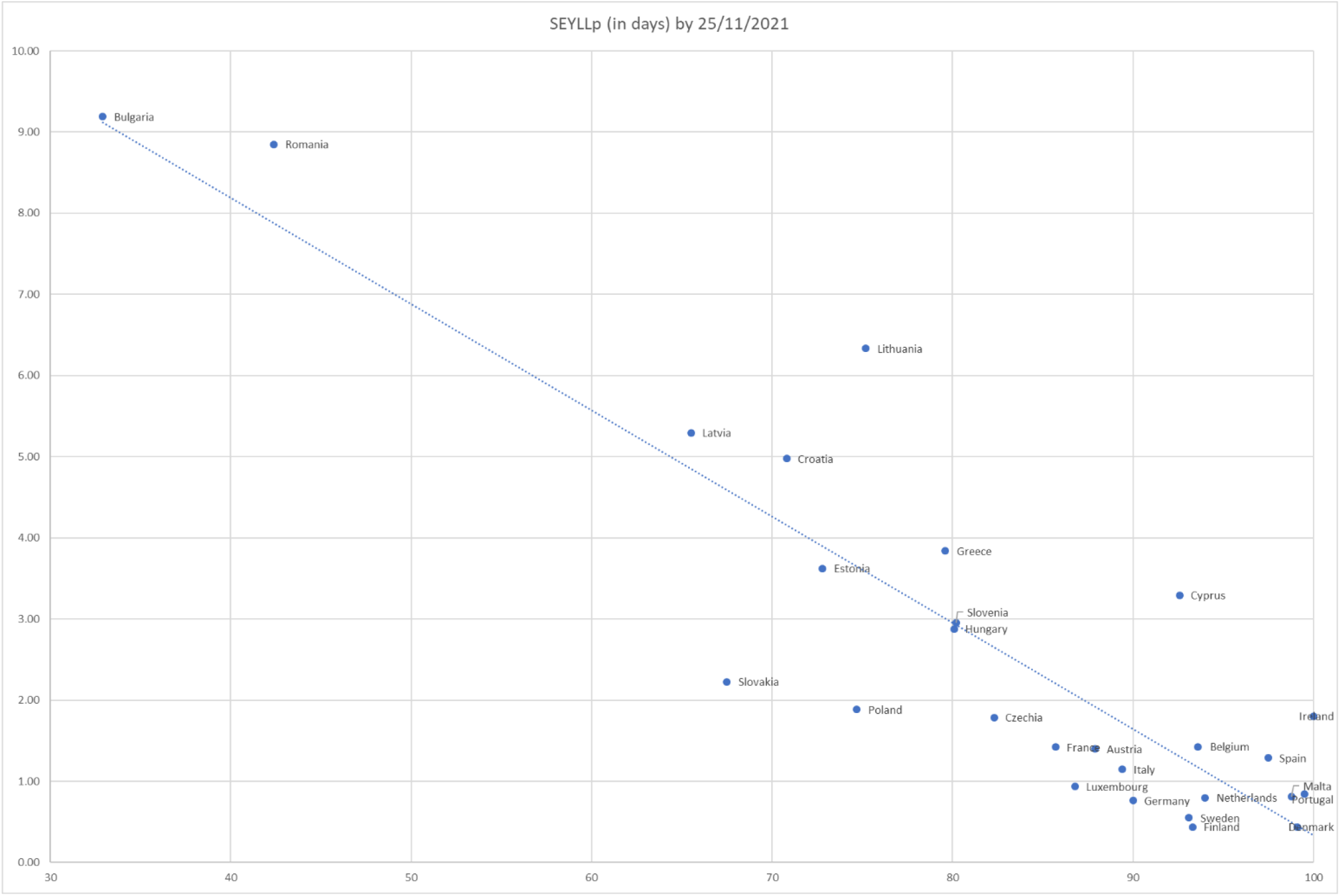
SEYLL_p_ (in days) for the population over 60 in the 27 E.U. countries from 25/5/2021 to 25/11/2021 vs. their percentage vaccination coverage.

SHARE’s second iteration COVID-19 Survey vaccination intention percentages correspond well with E.U. country-level vaccination rates reported by the European Centre for Disease Prevention and Control for the 60+ populations by 25 November 2021 (ECDC vaccinetracker, 2021). The percentage of those over 60 who say they did not want to get vaccinated is used to approximate the *upper* vaccination limit in case of enforcing mandatory vaccination for these ages in every E.U. country. The actual vaccination percentages were used for the only two countries where vaccination percentage slightly exceeded this limit by 25 November 2021, namely Denmark and Portugal. Using these values as an input, the estimated SEYLL_p_ (in days) using the calculation approximation function is illustrated in Table 3. Figure 3 highlights the *differences* between the 27 E.U. countries’ SEYLL_p_ for the 60+ population by 25/11/2021 and their estimated respective values in the case of mandatory vaccination. In other words, Figure 3 shows the potential gains in fewer lives lost should mandates succeed to lead every recalcitrant individual to vaccination.

**Figure 3.**
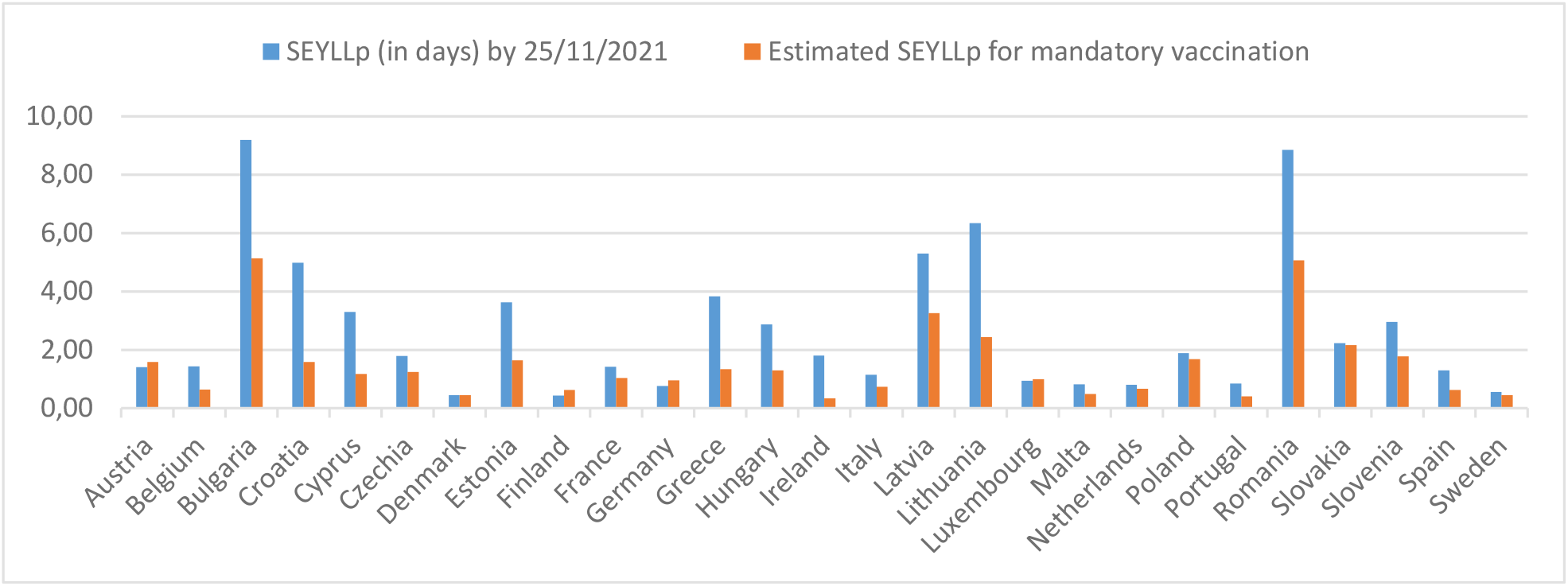
Differences between the 27 E.U. countries’ SEYLL_p_ for the 60+ population by 25/11/2021 and their estimated respective values in the case of a mandatory vaccination.

## Discussion

The simple calculations of this paper indicate that vaccination emerges to be the most important factor in explaining variations in in E.U. countries’ mortality from COVID-19 for age groups over 60. This result emerges naturally, without needing to employ complex mathematical models nor to bring other variables to bear. Its impact is significant, not only in terms of the number of deaths but also in terms of years of life lost that could be averted if the vaccination coverage in this age group increases.

More specifically, the values of yielded R^2^_adj_ and R^2^ , 79.62% and 78.8% respectively, and indicate that almost 80% of the variability COVID-19 mortality rates for the ages over 60 in E.U. countries can be explained solely from their vaccination coverage. In addition, the E.U.-wide approximate value function for SEYLL_p_ yields a Mean Absolute Deviation (MAD) of 0.82 days for the vaccination coverage on 25/11/2021, implying an impressive fit for such a parsimonious model. The number of deaths occurring at ages above 60 justifies policy responses to protect these vulnerable population groups. Whereas vaccine mandates exist and are common in all E.U. countries for children and childhood diseases, fully one year after the vaccination kick-off against COVID-19, there appears to be a reticence to proceed with equivalent mandates for COVID to affect the adult population. The *need* for such a mandate – in the sense of the share of the vulnerable groups who are not vaccinated voluntarily differs widely in the EU.

However, from Figure 3 it is obvious that in, say Bulgaria, Croatia, Cyprus, Estonia, Greece, Hungary, Latvia, Lithuania and Romania, even if the predicted upper vaccination limit is achieved and therefore mandates are 100% succesful, SEYLL_p_ would still remain high, as death rates are high. Thus, in order to bring the impact of the pandemic under control, the adaptation of some NPIs will still be necessary, which is aligned with the corresponding literature stream. Especially for Bulgaria and Romania, even when the lower bounds of the 95% CI are used for the estimation, there will still be a need for additional NPIs to minimize SEYLL_p_, as it will be greater than one day per semester.

The study has several limitations. As countries are at different stages of the pandemic trajectory, it is a snapshot of the impacts of COVID-19 on SEYLL_p_ by November 25, 2021. The Delta variant impacts in the examined time span of six months are probably mostly captured, but the advent of the Omicron variant at the end of November 2021 and its effects are not. The Omicron variant spreads more easily than the Delta and it is expected that anyone with Omicron infection can spread the virus to others, even if they are vaccinated or asymptomatic. Current vaccines are expected to protect against severe illness, hospitalizations, and deaths due to infection with the Omicron variant, but breakthrough infections in elderly people who are fully vaccinated are likely to occur, with an impact that necessitates more time to estimate.

Futhermore, SEYLL_p_ figures may be misestimated. On one hand, COVID-19 deaths may not be accurately recorded in some countries; most of the evidence suggests that there is a net underestimation of the total death toll on the aggregate level. On the other hand, those dying from COVID-19 may be an at-risk population whose remaining life expectancy is shorter than the average person’s due to co-morbidities (Devleesschauwer et al., 2020). Consequently, SEYLL_p_ due to COVID-19 may be overestimated.

Nevertheless, the study confirmed the large mortality impact of COVID-19 among the elderly, even using a metric that places greater weight on deaths occurring at younger ages. It also calls for devising policies that protect vulnerable demographics losing the largest number of life-years. Finally, in order to evaluate the effectiveness of mandatory vaccination against COVID-19 for ages over 60, country-specific data must be studied carefully, focusing on the vaccination adherence divergence in the E.U..

## Data Availability

All data produced in the present study are available upon reasonable request to the authors

